# Trust in, acceptance of, and responsibility for, artificial intelligence in healthcare – patient and healthcare practitioner considerations

**DOI:** 10.64898/2026.03.16.26348461

**Authors:** Emma-Jane Spencer, Imane Ihaddouchen, Stefan Buijsman, Jiwon Jung, Joris van der Vorst, Dirk Grünhagen, Kees Verhoef, Diederik Gommers, Michel van Genderen, Denise Hilling

**Affiliations:** Department of Adult Intensive Care, Erasmus MC, University Medical Center Rotterdam, Rotterdam, The Netherlands; Erasmus MC Datahub, Erasmus MC, University Medical Center Rotterdam, Rotterdam, The Netherlands; Faculty of Technology, Policy, and Management, Delft University of Technology, Delft, The Netherlands; Faculty of Industrial Design Engineering, Delft University of Technology, Delft, The Netherlands; Department of Surgical Oncology and Gastrointestinal Surgery, Erasmus MC Cancer Institute, University Medical Center Rotterdam, Rotterdam, The Netherlands

**Author notes:** **Corresponding author:** Dr. Denise Hilling, Erasmus MC Datahub, University Medical Center Rotterdam, Department of Adult Intensive Care (internal post address – Room Ne-403), Doctor Molewaterplein 40, 3015 GD Rotterdam, the Netherlands. These authors contributed equally to this work and share first authorship (Emma-Jane Spencer, Imane Ihaddouchen).

**Keywords:** Artificial intelligence, Healthcare, Applied ethics, Trust, Responsibility, Acceptance

## Abstract

**Objectives:** Using qualitative methods, this study aimed to provide a comparative overview of the similarities and differences in perspectives towards AI in healthcare in two different stakeholder groups: healthcare practitioners and patients. It also aimed to investigate whether these perspectives may influence the adoption of AI in healthcare.

**Design:** This study was conducted using semi-structured interviews. Qualitative data from the interviews were analyzed using both deductive and inductive coding, followed by a thematic analysis to identify the prevailing categories for further discussion.

**Setting:** The study was conducted within the Department of Surgical Oncology and Gastrointestinal Surgery of Erasmus Medical Center in Rotterdam, the Netherlands.

**Participants:** A total of 30 participants were recruited using purposive sampling based on predefined inclusion characteristics. This included 18 healthcare professionals (subdivided into 10 surgeons and 8 nurses), and 12 patients. The inclusion criterion for healthcare professionals included surgeons specializing in gastro-intestinal surgery, while the inclusion criterion for patients included those patients who had undergone gastro-intestinal surgery within the past 12 months at the time interviews were conducted. Exclusion criteria involved excluding patients with major health complications.

**Outcome measures:** The study’s central objective was to develop a set of thematic domains that characterize how both groups of stakeholders view the integration of AI in healthcare, encompassing their attitudes towards trust, acceptance, and responsibility. Additionally, it aimed to compare perspectives between healthcare professionals and patients in order to identify areas of convergence and divergence.

**Results:** The analysis comprised a total of 3 main thematic categories, with 10 subcategories. The main thematic categories which emerged were AI Knowledge, Ethics, and Operational and Clinical Implications. While clinicians largely focused on validation, monitoring, administrative labor, and clinical integration, patients emphasized the importance of human attention, of being heard, and of maintaining trust in their clinician.

**Conclusion:** Comparing the attitudes and perspectives of both healthcare practitioners and patients revealed the importance of taking into consideration both groups of stakeholders. While both groups tend to raise concerns about similar themes connected to responsibility, it is clear that this concern involves complex dynamics present in the epistemic environment of healthcare.

**Strengths and limitations:**

- This study uniquely compared healthcare practitioners’ and patients’ perspectives within a single qualitative design, using similar interview guides to enable direct cross-stakeholder comparison.
- The study examined perceptions of an AI model that had been designed and validated for clinical use, enhancing the practical relevance of the findings.
- The relatively small sample size may have limited the diversity of perspectives captured and reduced transferability.
- As the study was conducted in a single academic hospital in the Netherlands, the findings may not be generalizable to other healthcare settings or national contexts.

## Introduction

The proliferation of artificial intelligence (AI) as we now think of it has been on the rise since the early 2020s, with many scholars addressing the so-called “hype” narrative (Rose, 2025). This boom has affected all sectors of the economy to varying degrees, with predictions often made by experts about the possible impact it would have in years to come. Healthcare has been no exception. The possibilities AI could offer at the bedside have led to major investment in health-related AI research and development, in the hopes that reliable and trustworthy technologies would soon be implemented (Silicon Valley Bank, 2025). However, several recent studies have shown that, in spite of the focus and investment dedicated to AI, there remains a major implementation gap, with studies reporting that, in some domains of healthcare, only two percent of models make it to the bedside (Berkhout, 2025; Van De Sande, 2021). Furthermore, several years after the start of the intensive AI growth period, it is worth re-assessing the state of the art, asking whether or not sufficient progress has been made at the level of implementation to justify the substantial investment – of both focus and finance. In order to investigate the status quo, however, a simple overview of the operational AI systems in healthcare is insufficient. Instead, this approach must be combined with qualitative research into the views of those working in, and affected by, healthcare.

Against this background, there is a clear need for empirical research that examines how artificial intelligence is currently perceived by those directly involved in, and affected by, healthcare practice. This is particularly important in light of the fact that the patient perspective remains underrepresented in empirical studies. Thus, this study addresses this gap by offering a qualitative analysis of contemporary attitudes towards AI in healthcare, capturing perspectives from both healthcare professionals and patients. By comparing similarities and differences between these groups, the study provides insight into how AI is understood and evaluated across distinct yet interrelated stakeholder positions.

Beyond simply mapping attitudes, these findings are interpreted in light of broader ethical and epistemological considerations related to the operationalization of AI in clinical settings. In doing so, this study contributes to ongoing debates on AI integration in healthcare by foregrounding concerns that are frequently underrepresented in implementation-focused discussions, including questions of responsibility, knowledge and uncertainty, and the conditions under which AI systems are regarded as trustworthy. These questions are examined through the lens of ethical and epistemic dimensions for both physicians and their patients. Together, these contributions offer a nuanced assessment of the current moment in the adoption of AI in healthcare and help contextualize the persistent gap between technological development and clinical implementation.

## Methods

### Study design

An exploratory qualitative study design which employed semi-structured interviews with both healthcare professionals in the Department of Surgical Oncology and Gastrointestinal Surgery and patients at Erasmus Medical Center was used to explore attitudes towards AI in healthcare. Particular emphasis was placed on themes of trust, acceptance, and responsibility.

### Participants and setting

Participants were recruited using a purposive sampling strategy to capture a range of perspectives on the use of artificial intelligence in healthcare. Participants were selected based on their relevance to the research question and their ability to provide experiential insights into AI in clinical care.

Healthcare professionals, including nurses and surgeons working within the Department of Surgical Oncology and Gastrointestinal Surgery, were invited to participate via departmental email and were also approached in person to facilitate participation across professional roles. Patients were recruited from a gastro-intestinal surgery outpatient clinic through screening for eligibility by the treating surgeons. In line with ethical considerations, particular attention was paid to potential vulnerability during recruitment. Only patients who had undergone gastro-intestinal surgery within the preceding year were eligible to participate, ensuring that patient perspectives were informed by recent care experiences.

Recruitment continued until thematic saturation was reached within and across participant groups, defined as the point at which no new themes emerged from successive interviews.

Interviews with HCPs were conducted in person within the hospital setting, while the majority of patient interviews were conducted online using Microsoft Teams to facilitate participation.

### Data collection

Data were collected through semi-structured interviews, conducted by members of the research team with diverse academic and professional backgrounds, such as ethics, philosophy, and technical medicine. This interdisciplinary composition was intended to support reflexivity and enable exploration of both technical and normative aspects of artificial intelligence in healthcare.

A semi-structured interview guide was used to ensure consistency across interviews while allowing flexibility to explore topics raised by participants. The interview guide was developed through multiple iterative rounds by researchers with expertise spanning medical, technical, and social science disciplines. Additional methodological guidance was provided by an expert in qualitative research affiliated with TU Delft.

Interviews typically lasted between 30 and 45 minutes, although some were shorter and others much longer, depending on participants’ availability and depth of discussion. All interviews focused on participants’ experiences, perceptions, and expectations regarding the use of artificial intelligence in healthcare.

### Data analysis

Interview recordings were transcribed verbatim with internally developed software and were analysed using ATLAS.ti (version …) qualitative data analysis software. An initial preliminary codebook was developed collaboratively by the two first authors following familiarization with a subset of the transcripts. Both inductive and deductive coding was used to identity themes. To enhance analytical rigor, subsets of interview transcripts were then independently analyzed by the two first authors. The resulting codebooks were then compared and discussed to identify differences in interpretation and coding. Following this comparison, a shared codebook was fixed and the researchers revisited the full dataset, integrating additional codes and refinements derived from each other’s analyses. This iterative process allowed for continuous refinement of the codebook and supported the development of a coherent analytical framework that captured both shared and divergent perspectives across participant groups. Throughout the analysis, regular discussions were held to reflect on emerging themes and ensure consistency in coding and interpretation.

### Results

A total of 30 interviews were conducted, 18 of which were HCPs and 12 of which were patients. The researchers deemed it important to include nurses among the HCPs sample since they are the stakeholder group most responsible for the everyday care of patients, thus 8 of the HCPs were nurses and the remaining 10 were surgeons. The patient population tended to be older in age given the fact that many gastro-intestinal complications occur later in life.

## Ethics approval

This study involves human participants. The *Medisch Ethische Toetsings Commissie* (Medical Ethical Review Committee) (METC) at Erasmus Medical Center provided ethical approval for the study (Panama ID #13109). The researchers obtained informed written consent from every participant by explaining the purpose behind the study verbally as well as distributing an information letter. It was explained that all participation was entirely voluntary. Participants were informed that their confidentiality would be safeguarded and that interview transcripts would be anonymized prior to analysis to ensure their privacy. All collected data were secured using encrypted digital storage systems accessible only to authorized members of the research team.

## Results

The following themes emerged from the interviews with healthcare practitioners and patients. These themes were identified through both inductive and deductive means, iterative coding and comparative analysis across stakeholder groups. An overview of the themes and their corresponding subthemes is provided in Table 1.

**Table 1.** Emergent themes and subthemes from interviews with healthcare practitioners and patients, with accompanying operational definitions used by the research team.

| Theme | Subtheme | Definition |
| --- | --- | --- |
| AI Knowledge | AI Knowledge | Participants' awareness and understanding of what AI is, how it functions, its data inputs, validation, and perceived capabilities and limitations. |
| Ethics | Autonomy | Views on the impact of AI on patient decision-making, clinician judgement, and the preservation of autonomy and control. |
|  | Bias and fairness | Views regarding algorithmic bias, human bias, data representativeness, equity, and generalizability. |
|  | Doctor–patient relationship | Perceived effects of AI on communication, human touch, shared decision-making, trust between patient and HCP, and the relational dynamics of care. |
|  | Explainability | Expectations regarding the transparency, interpretability, and the ability to understand or justify AI-generated outputs – either to themselves or to others. |
|  | Responsibility and accountability | Perceptions of professional responsibility for decisions informed or supported by AI systems and their outputs. |
|  | Trust | Confidence in the reliability, safety, governance, and monitoring for the appropriate use of AI in clinical settings. The ability to rely on an AI model to assist with decision-making about patients. |
| Operational and Clinical | Clinical impact | Perceived effects of AI on clinical decision-making, patient outcomes, administrative efficiency, and quality of care. |
| **Implications** |  |  |
|  | Technical concerns | Practical issues related to system accuracy, integration, data quality, monitoring, and technical reliability and trustworthiness. |
|  | Workflow | Anticipated operationalization of AI in the clinical setting – in particular, how it pertains to efficiency, ease of integration. |

## AI knowledge

### AI knowledge

Across the interviews, both healthcare professionals and patients demonstrated a broad awareness for the fact that AI is increasingly visible in society and healthcare. However, the depth of understanding varied considerably. While some participants—particularly a subset of physicians—spoke in technical terms (eg, performance, validation), many described their AI knowledge as general, media-informed, or “surface-level.” Importantly, limited literacy did not necessarily imply rejection; instead, participants often framed knowledge as a condition for appropriate trust, safe use, and meaningful communication with patients.

Healthcare professionals commonly framed AI as a growing trend and described seeing it emerge across clinical domains (eg, imaging, diagnostics, prediction). This sense of momentum was echoed by patients, who often referred to AI in everyday contexts outside healthcare (eg, social media, synthetic content), suggesting AI has become a familiar societal phenomenon—though not always well understood.

> “The first thing is that it’s becoming more and more… it’s like really popular to do something with AI at the moment.” (HCP8)
>
> “There are already many applications in diagnostics… radiology, also pathology… identifying whether someone can possibly be discharged, or the occurrence of complications.” (HCP10)
>
> My husband has Facebook and I have Instagram, and you often see videos and photos there. At the beginning you could recognise from 10 km away that they were fake, but now you can’t see that anymore.” (PT10)

To some participants, this broader visibility of AI produced a sense of inevitability—AI as “the future”—while also raising awareness of how rapidly the technology is changing.

> “I think it’s kind of our future… we’re going to work with it everywhere anyway.” (HCP18)

A prominent pattern was uneven AI literacy. Several participants explicitly positioned themselves as lacking the knowledge to evaluate AI systems, sometimes describing AI as a vague umbrella term. This was seen among both healthcare professionals and patients, with patients often linking their limited confidence to low digital/technical familiarity.

> “Well, I know far too little about it, so AI is a catch-all term for me…” (HCP6)
>
> “No, I’m not at all up to date on that… I don’t know whether [robots in surgery] also has to do with AI.” (PT10)

In contrast, some physicians demonstrated more detailed reasoning, including concerns about imprecise “AI” labelling and the importance of quantifying model error.

> “Nowadays everything is called AI… sometimes it’s just a statistical model… I do think the terminology should remain precise.” (HCP1)
>
> “Even if it’s very good… then you have 10% error.” (HCP5)

Rather than focusing primarily on the technical workings of AI systems, many clinicians emphasised that what matters most is understanding limitations, including uncertainty, imperfect accuracy, and the strength of evidence behind a model. Several clinicians noted that predictive models inevitably produce errors and therefore require cautious interpretation in clinical practice.

> “There will of course be a certain percentage where the model is wrong. It will never be 100% water tight.” (HCP4)

Awareness of these limitations was also reflected by patients, who often described trust as fragile and shaped by experiences with medical technology and the consequences of errors—particularly when stakes are high.

> “An AI should then not look like: yes, well in 99% it always goes well. No, it is precisely that one percent… you must keep a close eye on that.” (PT4)

When asked what clinicians should know, healthcare professionals often articulated a minimum literacy threshold: not necessarily the technical mechanics, but understanding where data come from, development context, and external validation. Several framed this as analogous to understanding the limitations of other diagnostics—sufficient knowledge to interpret output responsibly.

> “He or she does have to have information about where the model was developed and how it was externally validated… [then] does not need to know in detail how the model is put together.” (HCP6)

Consistent with this, some participants explicitly called for structured education or guidance—especially to avoid blind trust in “sophisticated” systems and to support clinicians in discussing AI credibly with patients.

> “If you’re not aware of these issues… a lot of people trust the technology because it’s so complicated… that’s the danger… if you don’t know what the caveats are.” (HCP5)

Many patients linked information to reassurance and fairness: if AI influences decisions about their care, they wanted to be told and to receive explanations in accessible language, ideally integrated into existing preoperative information pathways. At the same time, patients differed in how much detail they wanted, suggesting that AI communication may need to be adaptable rather than “one-size-fits-all.”

> “I think just like what you are doing now: involve the patient—or at least involve society more in it.” (PT9)

## Ethics

### Autonomy

It is frequently argued in the philosophical literature that AI systems may risk undermining human autonomy in healthcare decision-making (Prince and Lim, 2025; Bjerring and Busch, 2021; Mittelstadt, 2019). In the interviews, autonomy emerged as a central concern for both healthcare practitioners and patients, though it was framed differently across groups.

Among healthcare practitioners, there was a strong emphasis on preserving professional judgement and maintaining a “human-in-the-loop.” Many doctors demonstrated awareness of automation bias and the risk of uncritical reliance on complex technologies. One practitioner noted:

> “If you’re not aware of these issues, then you just interpret the model and say, okay, this is fine and this has been made with a very sophisticated technology. I think a lot of people trust the technology because it’s so complicated and they can’t imagine what’s going on there when it’s being made. But that’s the danger, I think, if you don’t know what the caveats are, etc.” (HCP5)

Despite recognizing these risks, practitioners consistently positioned their own judgement as decisive. In discharge decisions, safety was described as overriding algorithmic advice:

> “Then with this model—because this is about earlier discharge—for me it’s a no-brainer to stay on the safe side. Then I think: you know, I just don’t fully trust it, screw your model, I keep the patient.” (HCP1)

Many clinicians, including nurses, referred to tacit knowledge—described as intuition, a “gut feeling,” or the “clinical eye”—as an essential aspect of care that could not be programmed:

> “But as a nurse you sometimes just have such a bad feeling… you just have the idea that something is going on, but you can’t really put your finger on it.” (HCP17)
>
> “That is the experience of doctors: the more experienced you are, the better that feeling is developed, because you actually trained yourself, just like the AI model is trained with a lot of information from previous patients.” (HCP3)

Some participants likened this experiential judgement to the training of AI models, suggesting that expertise develops through repeated exposure to cases. A minority explicitly warned against epistemic dependence:

> “…no, we shouldn’t become completely dependent on those models. I always find that a bit dangerous.” (HCP2)

Patients’ perspectives were more varied. While some were comfortable with a more paternalistic approach, others emphasized involvement and bodily autonomy, expressing concern that AI-supported recommendations might strengthen the physician’s position and silence their own voices:

> “The doctor says you may go home and I say well I don’t feel good at all. You should be able to say that I think.” (PT6)

Contrary to some practitioners’ assumptions, many patients linked knowledge to autonomy and wished to be informed if AI was used in decisions about their care. However, desired levels of involvement differed substantially. Concerns about overreliance were shared across groups, as one patient remarked:

> “I’m scared of it a little bit. Yeah. People don’t use their own brains anymore.” (PT1)

Overall, practitioners framed autonomy primarily in terms of safeguarding professional judgement, whereas patients more often associated autonomy with information, involvement, and the ability to voice disagreement. Although both groups recognized the risks of automation bias, the locus of concern differed.

### Responsibility and accountability

Healthcare practitioners were unequivocal in stating that ultimate responsibility for clinical decisions supported by AI remains with the doctor. Across interviews, clinicians emphasized that decision-support systems do not replace professional accountability and that the final judgement must rest with the treating physician. At the same time, many practitioners expressed substantive concerns about the systems themselves, including low or incomplete data quality, missing values, limited transparency, potential bias, and the possibility of error. This created a notable tension: while clinicians asserted ultimate responsibility, they also described reservations about the reliability and epistemic robustness of the tools informing their decisions. In this sense, they appeared prepared to assume responsibility for systems in which they did not express full confidence. Responsibility was also described as shaped by professional hierarchy. When asked how they would respond to an AI output they did not trust, one nurse reflected on the difficulty of challenging medical authority:

> “…as a new nurse, we also have a lot of new colleagues that are a bit reluctant to say anything to the doctor because the doctor is the man in charge or the woman in charge. So I think it depends on the nurse and her experience or his experience. So I think that’s a bit tricky.” (HCP14)

This suggests that, although formal responsibility was attributed to physicians, practical responsibility in responding to AI outputs may be mediated by experience and institutional dynamics.

Furthermore, when prompted to elaborate, several practitioners acknowledged that responsibility might extend beyond individual clinicians to other actors, including developers, hospital leadership, and those responsible for implementing the system. Thus, while the dominant narrative positioned the physician as ultimately accountable, participants’ reflections indicated a more distributed understanding of responsibility in cases of problematic AI outputs.

Patients spoke less extensively about responsibility. In general, they agreed that the doctor bears ultimate responsibility for clinical decisions, even when informed by AI. However, some patients demonstrated greater attentiveness to the shared and institutional dimensions of accountability, noting that decisions often involve multiple professionals and organizational structures. One patient explicitly recognized this complexity:

> “Well that is a complicated question… So to then blame the doctor, no. I think: no, as long as he listens well to the advice and simply determines himself: that man can go home, and if he then does not feel well at home, then he comes back.” (PT4)

Overall, while both groups formally located responsibility with the physician, healthcare practitioners emphasized professional accountability within hierarchical structures, whereas some patients more readily acknowledged the distributed nature of responsibility across clinicians, institutions, and system developers.

### Explainability

Explainability emerged as a key consideration in participants’ reflections on the use of AI in clinical decision-making. While both healthcare practitioners and patients expressed interest in understanding how AI systems inform care, their expectations and concerns differed.

Among healthcare practitioners, most participants emphasized the importance of transparency regarding how models are trained and what variables they rely on. Clinicians frequently noted that understanding the inputs and limitations of a model would allow them to better judge when its outputs should be trusted or treated with caution. As one practitioner explained:

> “I just like knowing what’s in it, mainly because then you can also see what’s not in it—what might make it a different patient.” (HCP1)

At the same time, several practitioners acknowledged practical limits to explainability. Some noted that complex machine-learning systems may function as “black boxes,” making it difficult for clinicians to fully understand how outputs are generated:

> “And I wonder if it will be feasible for us to… really see through the model, how it’s built, so that we know what we are using.” (HCP9)

In light of these constraints, many clinicians emphasized the practical interpretability of outputs rather than full transparency of the underlying algorithm. Participants responding to the discharge model described valuing clear, actionable recommendations—such as whether a patient could be discharged—over detailed insight into the model’s internal logic:

> “Well, you basically have all the things here. Look, I was reading through it, and then I think: well, I don’t need this—I can discharge him.” (HCP1)

Similarly, some practitioners suggested that detailed explanations might not be necessary for every output or every patient, particularly if the model’s recommendations aligned with clinical expectations.

Patients approached explainability from a somewhat different perspective. None explicitly raised concerns about the “black box” nature of AI systems, which may reflect either limited familiarity with technical aspects of AI or a lack of perceived importance of this issue. However, many patients nevertheless expressed interest in receiving more information about how AI is used in their care, including both its role in decision-making and, in some cases, aspects of how the technology works.

This interest contrasted with assumptions expressed by some healthcare practitioners, who believed patients would not be concerned with such information. One practitioner stated:

> “No, patients are not busy with that at all. They are busy with very different things than looking at such a model.” (HCP11)

When asked if patients might require information about how decisions were reached using the model, another HCP laughed and said,

> “Patients just don’t do that.” (HCP1)

In contrast, several patients indicated that they would welcome greater transparency regarding the use of AI in clinical decision-making. When asked whether additional information about the model would be desirable, one patient responded:

> “Yes I would have found that interesting.” (PT6)

Taken together, these findings suggest that while clinicians primarily value explainability insofar as it supports safe and practical decision-making, many patients associate explanation with transparency and informational inclusion. The discrepancy between practitioners’ assumptions and patients’ expressed interest highlights a potential gap in expectations regarding how AI-supported decisions should be communicated in clinical settings.

### Bias and fairness

Participants were asked several questions connected to bias and fairness. While both healthcare practitioners and patients acknowledged the possibility of bias in algorithmic systems, the depth and framing of these concerns differed across groups.

Among healthcare practitioners, awareness of bias in medical data and algorithms was relatively widespread and, in several cases, exceeded the researchers’ expectations. Many clinicians drew on their familiarity with broader discussions in medical research, where issues of representation in clinical studies are well documented. One practitioner noted:

> “I think, well, if you look at it broader, you already read a lot about the research being done, especially in white men, compared to females, minorities.” (HCP7)

This awareness often translated into a cautious stance toward AI systems. Several clinicians emphasised the importance of critically assessing the populations on which models are trained and whether these correspond to the patients encountered in their own clinical context:

> “You obviously have to keep thinking: who were these data trained on? Who is included? And are those the patients I’m dealing with? I think that matters, because you can have a very different composition. I wouldn’t just believe it: if I worked in Japan, an AI system developed on Americans—I wouldn’t just apply it to my patient care.” (HCP1)

Despite this awareness, the degree to which bias was perceived as a major concern varied considerably. Some participants expressed significant worry about the implications of biased outputs for vulnerable patient groups, whereas others suggested that the operational benefits of AI might outweigh these concerns in practice. One practitioner candidly remarked:

> “…you don’t always want it, but it’s a matter of personal gain. Yeah, so if there’s a lot of personal gain in it but you’re not really sure if the model is really treating everybody equally, then yeah, that worry might be a little bit less if it saves you three hours a day. Yeah, that’s the honest answer.” (HCP9)

Several clinicians also reflected on the broader context of human decision-making, noting that bias is not unique to AI systems but is already present in clinical practice:

> “But bias is not specific to AI, you know.” (HCP6)

In many cases, concerns about bias were ultimately framed as issues of generalisability. Practitioners frequently questioned whether models trained in one population or healthcare system would perform reliably in other clinical settings.

Among patients, bias and fairness were discussed less frequently. Many participants appeared unfamiliar with the concept of algorithmic bias or had not previously encountered debates surrounding it. Nevertheless, a small number of patients connected the issue to concerns about whether certain social or demographic groups might be disadvantaged by the technology. One participant reflected on the possibility that their own background could influence how a system performs:

> “Yes, well… that can happen, yes. I am also of Moroccan origin, so yes, then I hear it too.” (PT3)

Overall, while healthcare practitioners tended to frame bias as a technical and methodological issue related to training data and generalisability, patients who raised the issue were more likely to interpret it in terms of potential impacts on particular social groups.

### Doctor-patient relationship

The potential impact of AI on the doctor–patient relationship generated divergent views among healthcare practitioners. Participants were divided on whether AI would affect this relationship at all. Some believed it would have little to no impact, others suggested it could strengthen trust by supporting clinical decisions with additional evidence, while a third group worried it might weaken the interpersonal aspects of care. Overall, there was little consensus among practitioners.

Despite these differences, most healthcare practitioners agreed that patients should generally be informed if AI is used in their care. Even when participants did not frame this as a strict legal requirement, many stated they would likely disclose this information to their patients in any case.

There was much discussion about the extent to which HCPs could try to convince their patients – either that AI was safe and ought to be used, or as evidence that they were safe to be discharged from hospital, in the case of the DESIRE model. One surgeon said,

> *“Yeah, well, it’s always about people are skeptical. At home I’m always skeptical when we have a new sofa or something like that. I say I hate it and in three months I actually really like it, so we have the right explanation. And if patients refuse, I would also give them the possibility that if they are really skeptical and I can actually try to explain the advantage and the benefits, but if they remain skeptical and they don’t want it, they could also have the choice that we don’t use it*.*”* <u>(HCP8</u>)

Other practitioners viewed AI models as potentially strengthening their clinical recommendations, particularly when patients were hesitant about discharge decisions. In these cases, the model could serve as additional evidence supporting a course of action:

> “You could also think the other way around: you’d use it more often when people are hesitating. That you say: well, we have a calculation model with hundreds of thousands of patients that really shows it’s safe to go home—then it even gives an extra little push. I’d use it like that.” (HCP1)

Similarly, another clinician noted:

> “But if you have this model, you can just say, well, see these six points, it’s probably best that you go home today because you fulfilled our metacriteria. Yeah. Yeah, why not?” (HCP8)

Among patients, the doctor–patient relationship was discussed somewhat less frequently than expected. However, when it was raised, participants often emphasised the importance of the human and relational aspects of care. Several patients expressed concern that AI systems might not capture subjective or embodied aspects of illness that clinicians recognise through direct interaction:

> “But he doesn’t know how you feel. He doesn’t have feelings. Every person has the feeling about his body and how does it feel, how is your body feeling. I don’t think a feeling is something you can measure in an AI?” (PT1)

Others highlighted elements of motivation, effort, and personal circumstances that they believed would be difficult for an algorithm to capture:

> “…I wanted to go earlier home… And so I managed to go home very quickly and with the model I think it doesn’t work. The model doesn’t see that you are very willing and intending to go home as quick as possible.” (PT7)

A related concern was the possibility that patients might feel unheard if clinical decisions relied heavily on algorithmic recommendations. One participant described this fear as central:

> “My fear would constantly be: are they really looking at me? That would be the core of my fear… But it would mainly be the personal part, how I feel and whether that is heard.” (PT5)

At the same time, several patients acknowledged that AI could potentially support better clinical outcomes and therefore did not reject its use outright. Instead, their reflections suggested that maintaining the interpersonal and attentive aspects of care would remain essential if AI systems are integrated into clinical practice.

### Trust

Trust played an important role in shaping participants’ willingness to accept the use of AI in clinical decision-making. Among healthcare practitioners, however, trust was rarely framed as a general attitude toward AI itself. Instead, clinicians emphasised the importance of empirical validation and continued oversight before such systems could be fully relied upon.

Many practitioners indicated that their trust would depend on seeing evidence that a model performs reliably in practice. Participants frequently emphasised the need for both initial validation and ongoing monitoring once systems are implemented. Several clinicians also expressed interest in being involved in this monitoring process, reflecting a desire to maintain active oversight of algorithmic outputs:

> “…so what were my factors that I included in the decision that’s probably not integrated into the model? And then talk to the developers and see how they can improve.” (HCP5)

Interestingly, discussions of trust among healthcare practitioners often centred less on trusting the AI system itself and more on the way such systems might support clinicians’ own confidence in decision-making. Several participants described AI as providing reassurance or confirmation for decisions they were already inclined to make:

> “Then this gives that push in the back, as support.” (HCP1)

At the same time, some clinicians recognised that increased trust in the system could lead to greater reliance on its outputs:

> “So the more trust, the more you’ll rely on it, I think.” (HCP1)

Among patients, attitudes toward trust followed a different pattern. While participants did not generally express strong distrust of AI, they also did not frame trust primarily in relation to the technology itself. Instead, patients consistently emphasised that their trust lies with their treating physician, whom they expect to use available tools responsibly in making clinical decisions.

One participant described this relationship as follows:

> “I’ve also always had the idea of I’ll listen to the man in charge or the woman in charge or the doctor in charge. Ultimately, he makes the decision. They say, you know, it’s fine for you to go home. And I say, thank you very much. I will. And if they say, you know, it’s absolutely necessary you stay another two days or a week in the hospital. That is what I’ll do. So I’m a bit of a follower.” (PT11)

In this sense, patients tended to view AI as part of the broader set of tools available to clinicians rather than as an independent source of authority. Their trust in the technology was therefore mediated through their trust in the physician responsible for their care.

## Operational and clinical implications

### Perceived clinical impact of AI

Across interviews, both healthcare professionals and patients expressed a generally positive view of AI in healthcare. Many participants believed that AI systems could support clinicians by processing large amounts of data and identifying patterns that may be difficult for humans to detect. Several clinicians suggested that AI might complement clinical experience by drawing on information derived from large patient populations.

> “A beginning doctor has maybe seen fifty patients and has to base their feeling on that. And now they have a tool that is trained on thousands of patients.” (HCP3)

Participants also linked the potential benefits of AI to broader structural challenges in healthcare, such as workforce shortages and increasing complexity of care. Some patients believed that AI could help make healthcare more efficient and allow clinicians to focus more on direct patient care.

> “Healthcare is difficult to organise. There’s a shortage of people. If AI can help with some tasks so that staff have more time for patients, then I think it’s a very good idea.” (PT12)

Despite this general optimism, several clinicians expressed uncertainty about the actual added value of AI systems in clinical practice. Some participants noted that many AI initiatives promise substantial improvements, yet tangible benefits in everyday care remain limited. This led some clinicians to adopt a “wait-and-see” attitude toward new AI applications.

> “We’re now in a situation where there’s a lot coming that is supposed to improve things—but we’re not really seeing that yet. So it’s a bit ‘first see, then believe.” (HCP1)

Others emphasised that the development of AI tools should begin with a clearly defined clinical problem, rather than assuming that AI itself will provide the solution.

> “You can start collecting anything and say it’s interesting, but you should first ask: do we really need it and does it actually solve the problem we want to address?” (HCP8)

When discussing the discharge model specifically, several clinicians described how a predictive model could function as a form of decision support. Participants emphasised that such a tool would not replace clinical judgement but could serve as an additional source of information to support decision-making.

> “I am generally positive about it. It can be supportive to what we now do, which is still largely based on experience and feeling. It could add something to what we already do.” (HCP10)

Some clinicians suggested that the model could help highlight patients who may be ready for discharge or provide additional reassurance when discussing discharge decisions with colleagues.

> “When you discuss with colleagues whether a patient can be discharged, showing that the model predicts a good probability of safe discharge could really help.” (HCP8)

Others noted that the model might also have educational value by highlighting which clinical variables contribute to discharge readiness.

A recurring theme was the potential role of the model in situations where clinicians experience uncertainty. Several healthcare professionals explained that discharge decisions often involve incomplete information and clinical judgement, particularly when balancing patient safety against hospital capacity.

> “In doubtful cases it could be decisive, because you don’t always have everything in mind. The program might know the whole file and give that final push.” (HCP13)

Participants also suggested that the model could function as a neutral reference point in discussions between clinicians, for example when nurses and physicians have different perspectives on whether a patient is ready to go home.

> “Sometimes nurses and doctors see things differently about discharge. A model like this could help by saying: based on these factors the patient can probably go home.” (HCP14)

Some participants believed that the model could contribute to improved patient outcomes by identifying patients who could safely leave the hospital earlier or by flagging those who may require additional monitoring or aftercare.

> “We probably have quite a lot of patients who stay in the hospital longer than necessary.” (HCP7)

Patients also associated such systems with potential improvements in efficiency and access to care, suggesting that earlier discharge could free up resources for other patients.

At the same time, participants highlighted potential risks associated with AI-supported discharge decisions. The most frequently mentioned concern was the possibility that patients might be discharged too early, potentially leading to complications or readmissions.

> “If you discharge people too early, they may come back through the emergency department. Then you have a higher readmission risk.” (HCP1)

Patients similarly emphasised that recovery trajectories differ between individuals and that clinical decisions should remain sensitive to individual circumstances.

> “Every patient is different. One recovers faster than another, and that’s difficult to capture completely in a system like that.” (PT5)

However, some clinicians suggested that the potential negative consequences of errors in this specific application would likely be limited compared with other medical decisions. In their view, clinicians would remain able to override the model if necessary.

> “If I don’t agree with the advice, I’ll stay on the safe side. The worst that can happen is that the patient stays in the hospital two days longer—yeah, so what?” (HCP1)
>
> “It can’t really do any harm—it can only help, I think. Because you still have to keep thinking yourself.” (HCP17)

Patients also compared the potential risks of this application with more severe medical errors, suggesting that the consequences of an incorrect discharge prediction would likely be relatively limited.

> “It’s different if you end up in hospital for something and they amputate your leg when it wasn’t necessary—that’s when you can say something really went wrong.” (PT10)

These perspectives indicate that participants primarily viewed the model as a supportive tool, with potential risks perceived as relatively limited within this specific clinical context.

### Integration into clinical workflow

Many participants believed that AI systems could help reduce workload in clinical practice. Clinicians frequently linked this expectation to the increasing administrative burden associated with electronic health records and documentation requirements. Several participants expressed hope that AI could automate tasks such as writing reports or retrieving relevant patient information, allowing clinicians to spend more time on direct patient care.

> “I’d rather look at the patient than at the computer. If the computer can do something for me that saves time, I think that’s very good.” (HCP2)

Others noted that AI systems could rapidly process and interpret large amounts of clinical data that clinicians would otherwise need to manually review.

> “All the data that I would normally have available but would have to look up and interpret myself—an AI model can process that in half a second.” (HCP3)

Participants also linked the potential efficiency gains of AI to broader workforce challenges in healthcare.

> “Healthcare is difficult to organise. There’s a shortage of people. If AI can help with some tasks so that staff can focus on the important work at the bedside, then I think that’s a very good idea.” (PT12)

When discussing the proposed discharge model specifically, some participants suggested that such a tool could streamline decision-making and allow clinicians to work more efficiently.

> “If the doctor makes that decision with the help of the AI model, which has access to a lot of data, then that’s fantastic—because it means they can work faster.” (PT2)

Despite this optimism, many healthcare professionals emphasised that the success of such tools would depend heavily on how they are integrated into existing clinical systems. Participants stressed that the model should operate seamlessly within the electronic health record and require minimal additional effort from clinicians.

> “The condition for this to succeed is that the advice just appears somewhere. If you have to tick a hundred boxes to get it, then it becomes just another tool you have to work for.” (HCP1)

Similarly, participants emphasised that the model should be accessible directly within existing clinical software rather than requiring clinicians to open separate systems.

> “It should be available very easily within the patient’s chart… not through other portals. I don’t think people are going to open extra systems for every patient.” (HCP9)

At the same time, several clinicians expressed concern that AI tools could inadvertently increase workload rather than reduce it. Some participants reflected on previous digital innovations in healthcare that had been introduced with promises of efficiency but ultimately resulted in additional administrative tasks.

“Every improvement that has been presented as an improvement has meant extra work for the medical specialist.” (HCP1)

Others worried that clinicians might be required to review or verify multiple models or outputs, which could undermine the intended efficiency benefits.

> “If I have to click a box saying I know the model and its limitations, then when am I supposed to study all those models? That’s not going to save me time.” (HCP9)

Finally, participants raised practical questions about how the model would be incorporated into existing clinical decision-making processes. Some clinicians wondered how the model’s output should be documented in the patient record and how responsibility for decisions would be recorded.

> “How do you document this? Do you write in the file: ‘the model said discharge and I didn’t have an opinion, so I sent the patient home’?” (HCP5)

Others reflected more broadly on how the model would be used during clinical discussions about patient discharge.

> “I’m very curious about the practical implementation: will we briefly discuss each patient and say ‘the model suggests discharge tomorrow—does everyone agree?’” (HCP2)

Taken together, these findings suggest that while participants recognised the potential of AI systems to support clinical workflow, their acceptance was strongly contingent on seamless integration into existing systems and the ability to use such tools without introducing additional complexity or administrative burden.

### Technical concerns

Concerns about data quality were frequently raised when discussing the technical feasibility of AI systems in healthcare. Several clinicians emphasised that the reliability of AI models ultimately depends on the quality and completeness of the data on which they are trained.

> “The potential value of artificial intelligence really depends on the quality of the data. And that’s a big issue that we have not resolved yet.” (HCP5)

Participants also noted that the reliability of models may vary depending on the frequency with which certain clinical conditions occur in the available data. When only a limited number of cases are available, the resulting predictions may be less reliable.

> “Some procedures we perform very often and others hardly at all. So how reliable is your data then if you only see a condition a few times per year?” (HCP2)

Participants also expressed concerns about the development and validation of AI models. Several clinicians emphasised that prediction models should be rigorously evaluated before being implemented in clinical practice.

> “You really have to study whether it works that well. I certainly wouldn’t adopt it blindly, because there are too many variations that you don’t have insight into.” (HCP1)

In addition to initial validation, participants stressed the importance of ongoing monitoring after implementation to ensure that models continue to perform as intended.

> “You have to continuously check whether your model still functions as it should. You can’t just make a model once and then be done.” (HCP6)

Some clinicians also highlighted the importance of the population on which a model is trained, noting that models developed in one setting may not automatically generalise to another.

> “You expect that applicability might be greater if a model is developed in the Dutch population.” (HCP10)

Several participants raised concerns about the technical infrastructure required to support AI systems in clinical practice. In particular, clinicians pointed to challenges related to extracting and integrating data from electronic health record systems.

> “We tried automatic extraction from electronic patient records with AI, but it worked for one hospital and then failed for another. So getting this to work on a national level is quite difficult.” (HCP5)

Participants also emphasised that changes in hospital information systems could influence model performance, requiring continuous monitoring and maintenance.

> “Changes in the electronic patient record can also affect the model. You have to stay continuously alert to that.” (HCP6)

Finally, some participants questioned whether the variables included in predictive models would adequately capture the complexity of clinical decision-making. Clinicians emphasised that discharge decisions often involve nuanced clinical factors that may be difficult to represent in structured data.

> “Pain is very important after surgery, and also how someone is eating or mobilising. Those are all things you need to see before someone can go home.” (HCP16)

Patients similarly reflected on the difficulty of capturing individual recovery trajectories within predictive models.

> “Many things were already good in my case—my heart rate and blood pressure recovered quickly— but still I wasn’t ready to go home.” (PT5)

Taken together, these findings suggest that participants perceived several technical challenges related to the development, implementation, and maintenance of AI systems in healthcare, particularly regarding data quality, model validation, infrastructure, and the representation of complex clinical realities.

## Discussion

This study explored the different views from two stakeholder groups, patients and healthcare professionals, through the lens of DESIRE, an AI-based system designed to assist with post-operative discharge decisions. Participants showed broad awareness of AI in healthcare, but literacy varied from superficial, media-informed understanding to detailed reasoning among some physicians.

Limited knowledge did not equate to rejection; rather, participants viewed understanding as essential for safe use and trust. Clinicians emphasised understanding a model’s development context, data sources, and validation, echoing prior findings that AI literacy supports appropriate reliance and risk mitigation.[ref] Patients similarly highlighted the importance of transparent communication to support agency and informed engagement. Uneven literacy—particularly among non⍰technical clinicians and patients—highlights the need for targeted AI education. Structured training initiatives that focus on how AI works in clinical contexts and its limitations could mitigate risks associated with automation bias and inappropriate reliance on AI outputs [Schubert et al., 2025].

Under the theme of ethics, our findings suggest that the responsible use of AI in medicine unfortunately cannot be captured by guidelines or AI governance alone. Furthermore, while similar thematic issues arose for both HCPs and patients, their views regularly diverged from one another. Particularly within the patient group, it is difficult to pin down any consensus, and it appears that a one-size-fits-all approach would be insufficient to inform HCPs on how to incorporate AI into their clinical practice. Different patients require (and desire) different things.

As concerns the epistemic environment in which clinical decisions are made, generally there appeared to be a high requirement for participation in AI discussions, decision-making, and system monitoring. This was often true of patients as well as HCPs, which might surprise some of the HCPs.

HCPs did not express a general distrust toward AI systems, although they repeatedly emphasized the importance of validation, transparency about model logic, and ongoing monitoring in order to feel safe about AI use in their clinical environment. On the other hand, for patients, trust was not about AI systems themselves; instead, patients largely located trust in their clinicians, or in the hospital at large. We take this to be an indication of the importance of the doctor-patient relationship, particularly as AI is integrated into care pathways.

Regarding the kind of knowledge HCPs took AI outputs to be, it is concerning that so many HCPs spoke of AI support as a kind of “evidence” to convince patients of a particular course of action. AI outputs tended to be thought of as an additional tool on the side of the physician, potentially to be employed in cases where patients were not convinced of particular clinical decisions. On the other hand, many patients expressed worry or fear at the possibility of not being heard, something which has been flagged already in the literature on epistemic injustice (Emah and Bennett, 2025). This dynamic should flag as a deep concern and efforts must be taken to ensure that patients are not silenced by a power dynamic which attributes less clout to their voices in situations where AI “sides” with a physician.

Participants recognised the potential of AI to improve clinical care, particularly by supporting decision-making and enhancing efficiency. Clinicians emphasised that AI could synthesise large volumes of data, highlight patients ready for discharge, and provide reassurance in uncertain cases, aligning with literature showing that perceived utility is a key driver of adoption in healthcare settings [Zheng et al., 2025]. Participants also noted potential risks, such as premature discharge or reliance on incomplete model outputs, but emphasised that clinical judgement would remain decisive, consistent with prior work on human–AI collaboration [Wekenborg, 2025].

Integration into workflow was repeatedly highlighted as essential; tools must be seamlessly embedded in electronic health records and require minimal additional effort, echoing evidence that usability and alignment with existing processes strongly influence adoption [Prakash, 2021; Esmaeilzadeh, 2024]. Technical considerations—including data quality, local applicability, model validation, and infrastructure—were also raised, reflecting recognised barriers to safe and reliable AI deployment [Hassan et al., 2024].

Together, these findings suggest that the perceived clinical and operational benefits of AI depend not only on model accuracy but also on thoughtful integration, clear communication, and ongoing monitoring, highlighting the need for both organisational and technical preparation to support effective adoption.

## Conclusion

This study examined the perspectives of both healthcare practitioners and patients on the use of AI in clinical decision-making, highlighting the ethical, epistemic, and practical considerations relevant to AI systems such as the surgery discharge model, DESIRE. While clinicians emphasised the importance of validation, transparency, and maintaining oversight to avoid automation bias or excessive epistemic dependence, the inclusion of patient perspectives revealed additional concerns and expectations regarding autonomy, communication, and trust in the clinical relationship. In several instances, patient views challenged assumptions held by healthcare practitioners about what patients wish to know or how they may respond to the use of AI in their care. Taken together, these findings suggest that the responsible deployment of clinical AI systems depends not only on their technical performance but also on cultivating epistemic and ethical conditions that support clinicians’ critical engagement while remaining attentive to evolving patient expectations within the doctor-patient relationship. Considering both stakeholder perspectives is therefore essential for ensuring that AI contributes to ethically sound and medical practice.

## Data Availability

All data produced in the present study are available upon reasonable request to the authors

